# Examining the Influence of Displaced Populations on Tuberculosis Transmission: A Benue State Case Study

**DOI:** 10.64898/2026.10.20.26365996

**Authors:** Uduak Bassey, Temidayo Fawole, Thompson Ndifreke, Bethrand Odume, Ogoamaka Chukwuogo, Eze Chukwu, Chiedozie Abagwalatu, Ntamu Ntamu

## Abstract

**Background:** Tuberculosis (TB) remains a significant public health challenge, particularly among displaced populations. These environments foster TB transmission due to overcrowding, poor living conditions, and inadequate healthcare access

**Objectives:** To compare the proportion of tuberculosis (TB) cases detected through screening among displaced populations residing in internally displaced persons (IDP) and refugee settings with that among the general population in Benue State, Nigeria.

**Design:** Quantitative observational study comparing TB screening and diagnostic findings across displaced and general populations.

**Setting:** IDP camps, refugee settings and general-community screening locations in Benue State, Nigeria.

**Participants:** A total of 1,938 individuals were screened: 1,143 in displaced settings and 795 in the general population. Participants were assessed for presumptive TB and underwent the applicable diagnostic evaluation.

**Primary and secondary outcome measures:** The primary outcome was the proportion of screened individuals diagnosed with TB in displaced settings compared with the general population. Secondary outcomes were the numbers of presumptive TB cases and TB diagnoses classified as bacteriologically confirmed.

**Results:** Among the 1,143 individuals screened in displaced settings, 128 presumptive TB cases were identified, with 46 confirmed cases (26 bacteriologically and 20 clinically). In contrast, only 3 TB cases (2 bacteriologically and 1 clinically) were detected among 795 individuals in the general population. The TB prevalence rate in displaced populations was 4.0%, significantly higher than the 0.4% observed in the general population.

**Conclusions:** The study confirms a heightened TB burden in displaced communities, emphasizing the need for enhanced screening, diagnosis, and treatment strategies.

## INTRODUCTION

Tuberculosis (TB), an infectious disease caused by Mycobacterium tuberculosis, remains one of the leading public health challenges globally, particularly in low- and middle-income countries. Despite advances in TB control, regions affected by displacement, such as Nigeria and other conflict-affected countries, continue to experience disproportionately high TB burdens. Internally displaced persons (IDPs) and refugee populations living in overcrowded and unsanitary conditions face heightened risks of TB transmission (1,2). Studies have shown that displaced populations in IDP and refugee camps across Africa and South Asia experience higher TB prevalence due to limited healthcare access, poor living conditions, and nutritional deficiencies (3,4).

Nigeria, as one of the top 10 high TB burden countries, has faced increased TB transmission among displaced populations, particularly in states affected by insurgency and ethnic conflict. In Benue State, where violence has forced thousands into IDP and refugee camps, poor living conditions contribute significantly to TB outbreaks (5). Studies from several African settings have reported substantial TB burdens among displaced and refugee populations, with barriers to diagnosis and treatment complicating TB control in these settings (6-9). In the Democratic Republic of Congo, disrupted access to TB diagnosis and treatment has been reported among marginalized communities, highlighting the challenges of TB control in settings affected by poverty and limited healthcare access (10).

IDP camps provide conditions conducive to TB transmission, including inadequate ventilation, overcrowding, and malnutrition. Malnutrition weakens immune systems, making individuals more susceptible to TB infection and disease progression (11). Displaced persons often experience co-infections such as HIV, which further increases TB vulnerability (12). Studies in Sub-Saharan Africa highlight that TB-HIV co-infection remains a significant concern among displaced populations, leading to higher mortality rates and challenges in disease management (12-14).

Despite the growing TB burden in displaced populations, case detection and treatment remain major challenges. The World Health Organization (WHO) recommends Active Case Finding (ACF) in high-risk settings such as IDP camps to facilitate early detection and prevent further spread of the disease (1). However, security issues, stigma, and insufficient healthcare resources hinder the effective implementation of TB control programs in IDP and refugee settings (10,15). Stigma related to TB often delays diagnosis and treatment initiation, with affected individuals fearing social isolation or discrimination (16).

In addition to health consequences, TB significantly impacts the socioeconomic well-being of displaced individuals. TB morbidity and mortality lead to reduced economic productivity, increased healthcare expenditures, and prolonged displacement, further entrenching poverty in vulnerable communities (17). The economic consequences of TB in displaced populations have been widely documented, particularly in Nigeria and Sudan, where loss of income due to TB-related illness exacerbates financial instability (5,18). Moreover, TB outbreaks in refugee camps, such as those in Bangladesh hosting Rohingya refugees and in Lebanon among Syrian refugees, demonstrate that TB control among displaced populations is a global issue requiring urgent intervention (19,20).

Although studies have investigated TB prevalence and transmission dynamics in displaced populations globally, limited research has focused on insurgency-driven displacement in Nigeria. Most studies address TB in refugee camps in East Africa and South Asia, leaving a gap in understanding how displacement influences TB epidemiology in West Africa (21).

This study aims to address this gap by examining the impact of displacement on TB transmission in Benue State. Through a comparative analysis of TB prevalence in IDP camps and the general population, this research seeks to highlight the challenges faced by displaced communities and propose evidence-based strategies for improving TB control in these high-risk settings.

### METHODOLOGY

#### Study Design

This study employed an observational cross-sectional study that assessed the transmission of tuberculosis among displaced persons residing in insurgency-forced IDP and refugee camps in Benue State, Nigeria. It further included a comparison with the general population to provide some insight into how displacement influences TB prevalence. The main interventions of screening and the early detection of TB cases in these camps and the general population were done through an active case finding (ACF) strategy.

#### Study Sites and Population

Benue is a state in north-central Nigeria that has 277 wards and 23 Local Government Authorities (LGAs), (Figure 1). It has a total population of 6,015,633 projected from the 2006 national census. This makes Benue the 9th most populous state in Nigeria.

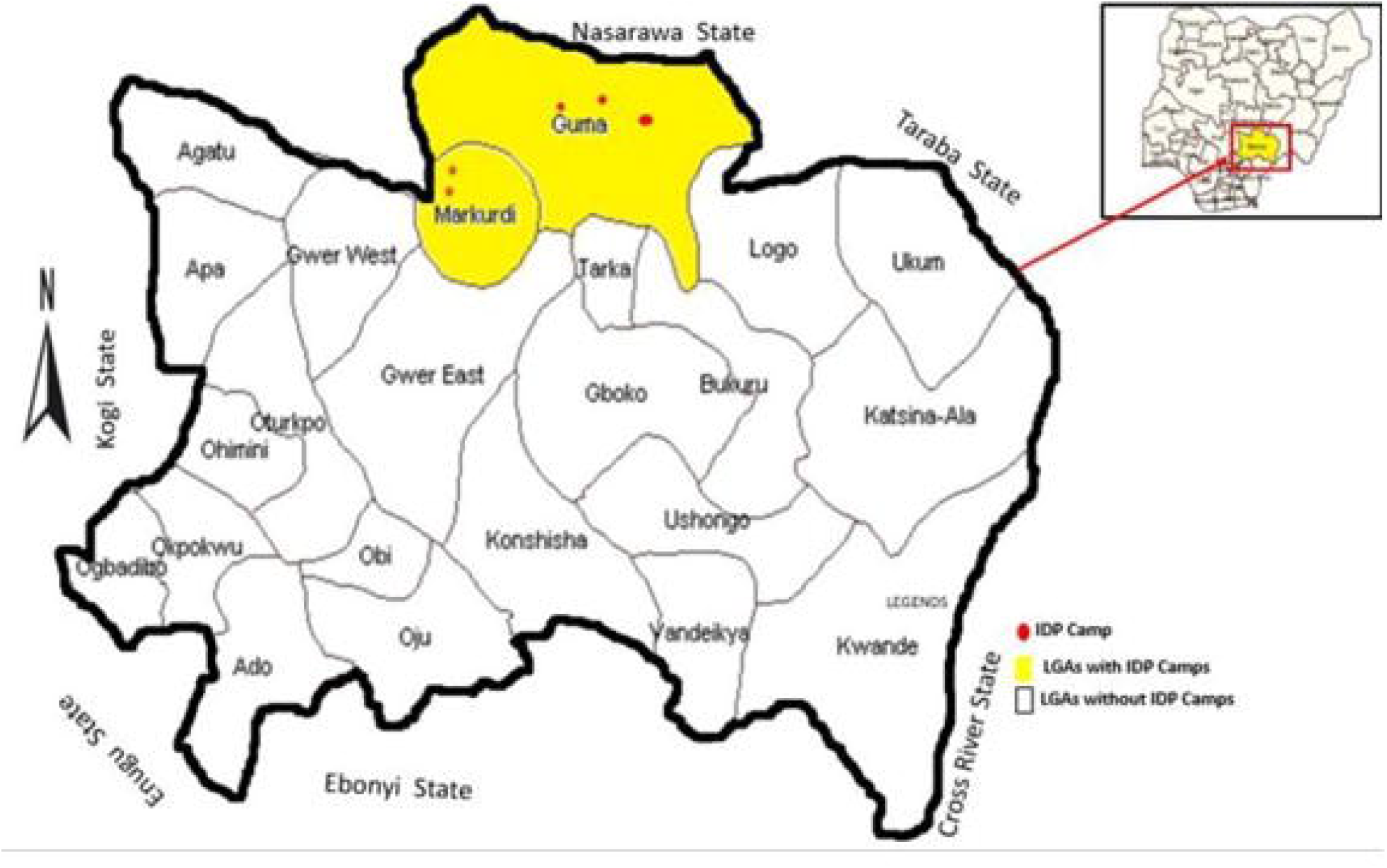

The places of study are three: two IDP camps in Guma LGA, one refugee camp in Kwande LGA, and the general population in Makurdi LGA of Benue State. This is selected based on the sizable population, proximity to conflict areas, and accessibility for healthcare interventions. The first IDP camp was screened from October 11-12, 2022, while the refugee camp was screened from October 3-4, 2023. The second IDP camp was screened between March 25-28, 2024. For comparison, the general population screening was conducted between March 11-15, 2024, in Makurdi LGA, a non-displaced Benue State community.

#### Data Collection Procedures

The study employed a multi-faceted approach to include clinical symptom screening, chest X-rays with artificial intelligence-based algorithms, sputum sample collection, and bacteriological testing. Prior to screening, community awareness campaigns were organized in each study site to increase participation and to inform the population about symptoms of TB and the importance of early detection.

#### CAD4TB Artificial Intelligence X-ray

Participants were screened for abnormalities indicative of TB using the CAD4TB Artificial Intelligence X-ray. Persons presenting abnormal chest X-ray results and participants reporting at least one of the common symptoms of TB, such as cough of more than two weeks, fever, night sweats, and weight loss, were considered presumptive TB cases. Sputum samples from all the presumptive cases were collected and taken to the laboratory for investigation of Mycobacterium tuberculosis using GeneXpert technology, which identifies also drug-resistant TB strains.

#### Sample Size and Screening

A total of 1,938 individuals were screened during the study. 0f these, 1,143 individuals were screened in the IDP and refugee camps, while 795 belonged to the general population.

#### Data Analysis

A quantitative analysis was conducted to compare TB prevalence among IDP camps, refugee camps, and the general population. Data was stratified based on location, number of screened individuals, presumptive TB cases, and bacteriologically confirmed TB cases. The calculation of TB yield in each study site for the analysis was also included to assess the difference in TB burden among displaced versus non-displaced populations.

#### Ethics approval

The activities described in this study were conducted as part of routine tuberculosis public-health and active case-finding services and were not initiated as a research study. The routine programme activities were therefore conducted without formal research ethics committee approval. Participants were informed about the screening procedures and provided verbal consent before screening and diagnostic testing.

### Patient and Public Involvement

Patients and members of the public were not involved in the design, conduct, reporting, or dissemination planning of this study. The study analysed data generated through tuberculosis screening activities.

## RESULTS

The breakdown of the study population and the number of presumptive TB cases identified is provided below.

A total of 1,938 individuals were screened across the IDP camps, refugee camps, and the general population. The breakdown of the study participants and the number of presumptive TB cases identified is presented in Table 1.

**Table 1:** Screening Cascade for all population screened Summary of TB Screening Results.

|  | **Number screened** | **Presumptive identified & evaluated** | **Bacteriological cases** | **Clinical cases** | **Number of patients on treatment** |
| --- | --- | --- | --- | --- | --- |
| Refugee | 300 | 23 | 10 | 1 | 11 |
| IDP 1 | 250 | 37 | 2 | 13 | 15 |
| IDP 2 | 593 | 68 | 14 | 6 | 20 |
| **Total** | **1143** | **128** | **26** | **20** | **46** |
| General Population | 795 | 28 | 2 | 1 | 3 |
| **Total** | **795** | **28** | **2** | **1** | **3** |

In the IDP and refugee camps, 1,143 individuals were screened, yielding 128 presumptive TB cases. After sputum analysis, 26 bacteriologically confirmed TB cases and 20 clinically diagnosed TB cases were recorded. In contrast, among the 795 individuals screened in the general population, only 3 TB cases were diagnosed, comprising 2 bacteriologically confirmed cases and 1 clinical case.

As shown in Figure 2, TB yield in the IDP and refugee camps was higher than that in the general population. Among the displaced populations (IDP and refugee camps), the TB prevalence rate was an aggregate of 4.0% (46 TB cases out of 1,143 screened), while the TB prevalence in the general population was 0.4% (3 cases out of 795 screened). However, across the IDP and refugee camps, the cases ranged from 3.6% to 6%, as shown in Fig 2.

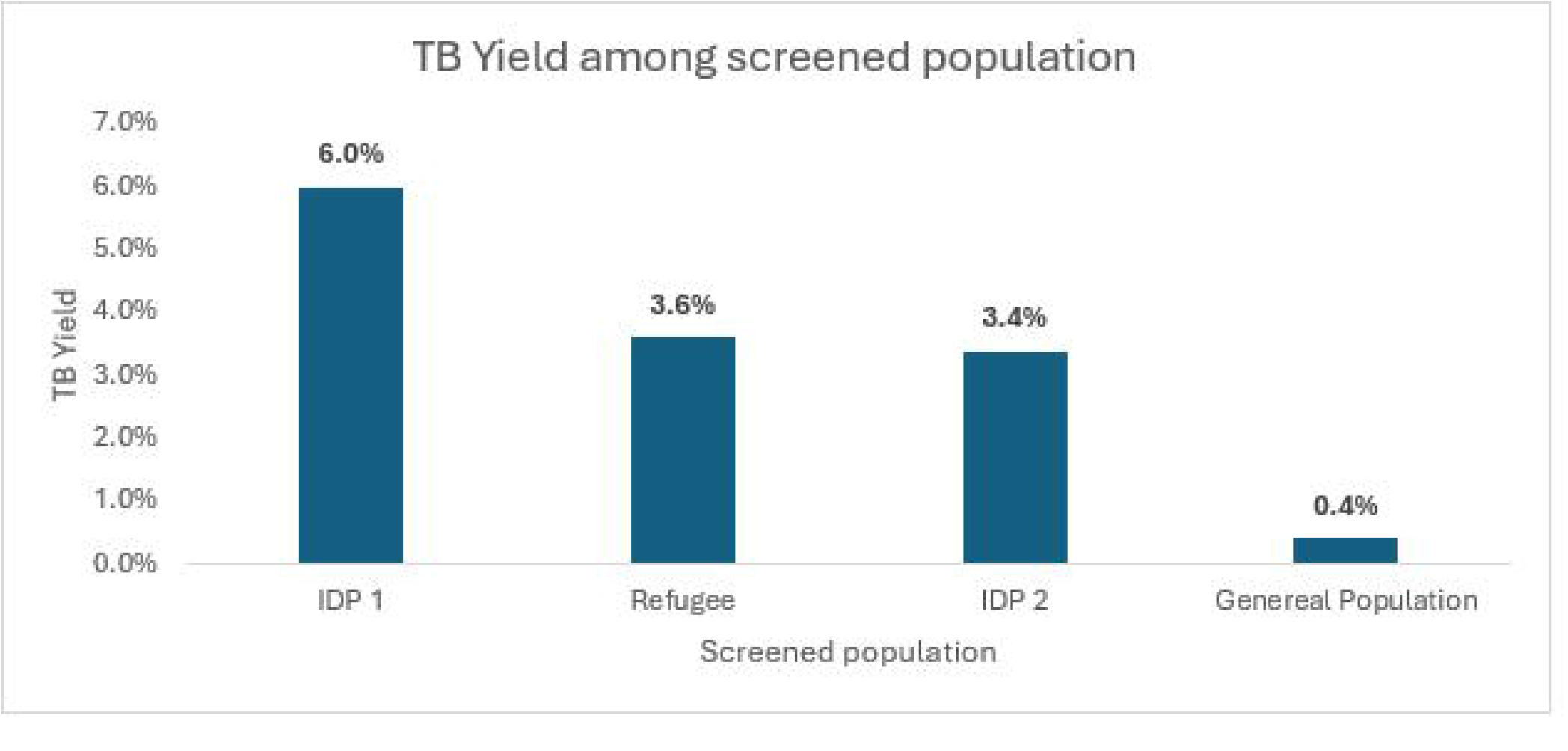

## Discussion

The results of this study confirm that TB prevalence is significantly higher among displaced populations compared to the general population, which aligns with findings from previous studies conducted in similar settings. Among the internally displaced persons (IDPs) and refugee populations screened, the TB prevalence rate was 4.0%, markedly higher than the 0.4% observed in the general population. The higher TB yield observed among displaced populations may reflect contextual factors commonly associated with displacement, including overcrowding, poor living conditions and barriers to healthcare access.

Findings from Ethiopia and Uganda similarly highlight that IDP and refugee camps act as TB transmission hotspots due to poor ventilation and inadequate healthcare services (6,7). A study by Bekele et al. (3) in Ethiopian refugee camps reported a TB prevalence rate of 3.9%, nearly identical to the findings in this study. Additionally, research from Kenya and South Sudan has demonstrated that TB prevalence in displaced populations is consistently higher than in surrounding host communities (4,6). These similarities reinforce the well-documented link between displacement and increased TB burden.

Several factors explain the elevated TB rates among IDPs and refugees. First, the high population density in camps facilitates airborne transmission of *M. tuberculosis*. Second, malnutrition and co-infections, particularly HIV, increase susceptibility to TB, as shown in studies across Sub-Saharan Africa (12-14). Additionally, delays in case detection due to limited diagnostic capacity and stigma further exacerbate TB transmission (15,16). Médecins Sans Frontières (11) has emphasized the challenges of delivering effective TB care in conflict settings, citing insecurity, healthcare worker shortages, and logistical constraints as major barriers to early diagnosis and treatment.

However, the TB prevalence observed in this study is lower than reports from certain conflict-affected regions such as South Sudan and the Democratic Republic of Congo, where TB prevalence among IDPs has exceeded 6% (7,10). One possible explanation is differences in screening strategies. This study employed active case finding (ACF), which has been shown to improve early TB detection and reduce transmission. In contrast, studies in South Sudan relied heavily on passive case finding, which often underestimates TB burden due to reliance on symptomatic individuals seeking healthcare services (13). The use of ACF may have led to the identification of more cases at an earlier stage, contributing to the lower prevalence observed in this study.

Additionally, TB prevalence variations may be influenced by differences in camp infrastructure and healthcare availability. In some regions, the presence of international organizations such as the WHO and non-governmental organizations (NGOs) has improved TB detection and treatment access, while in others, healthcare infrastructure remains severely underdeveloped (19,20). The findings of this study suggest that strengthening TB control efforts through targeted interventions, such as expanding ACF and improving diagnostic capacity in IDP camps, could significantly reduce TB transmission in these high-risk settings.

## Conclusion

These findings support the integration of systematic TB active case finding into health services for displaced populations in conflict-affected settings. The higher TB prevalence in IDP and refugee camps compared to the general population highlights the urgent need for enhanced TB screening, diagnosis, and treatment services in these settings. Lessons from similar studies suggest that implementing ACF, reducing stigma, and addressing comorbidities such as HIV could improve TB control efforts among displaced populations. Future research should focus on evaluating long-term TB treatment outcomes in displaced populations to better inform sustainable TB control strategies.

### Recommendation

Future research should explore the long-term effects of displacement on TB transmission, considering factors like migration patterns, healthcare accessibility, and nutrition. Assessing the effectiveness of mobile diagnostic units and molecular testing in resource-limited settings is crucial. Additionally, investigating TB-HIV co-infection and barriers to care, including stigma and healthcare infrastructure challenges, can help develop more effective, sustainable TB control strategies.

## Data Availability

All data produced in the present study are available upon reasonable request to the authors

## Acknowledgements

The authors acknowledge KNCV Nigeria, the National TB and Leprosy Control Program (NTBLCP), and field personnel involved in the routine active case-finding activities in Benue State. We appreciate their contributions to the implementation of these activities and the efforts of the personnel who supported TB screening and diagnostic services among displaced and general populations.

## Author Statement

None declared

## Author Statement

1. **Uduak Ikoedem Bassey:** Conceptualisation, data collection, interpretation of findings and drafting of the manuscript.
2. **Fawole Temidayo**: critical revision of the manuscript.
3. **Thompson Ndifreke:** Interpretation of findings
4. **Bethrand Odume:** Study design
5. **Chukwuogo Ogoamaka:** Supervision
6. **Eze Chukwu:** Supervision
7. **Ntamu Ntamu:** Data curation
8. **Abagwalatu Chiedozie:** Formal analysis

All authors: Reviewed and approved the final manuscript and agree to be accountable for the accuracy and integrity of the work.

## Data availability statement

The data supporting the findings of this study are available from the corresponding author on reasonable request, subject to applicable data access permissions and confidentiality requirements. The data are not publicly available.

## Article summary: Strengths and limitations of this study

### Strengths and limitations of this study

- The study used data from TB screening activities involving displaced populations and the general population in Benue State, Nigeria.
- Including a general-population comparison group provided a reference for examining differences in TB detection between the study populations.
- The study included individuals screened in real-world public health settings, supporting its relevance to TB case-finding activities.
- The findings may be affected by differences in participant selection, screening access and diagnostic evaluation between the populations, as well as the limited generalisability of findings from the selected locations.

## Strengths and limitations

A strength of this study is its use of TB screening data from displaced populations and the general population in Benue State, Nigeria, enabling comparison of TB detection across these settings. The classification of cases as bacteriologically confirmed or clinically diagnosed also provides a clearer description of the diagnostic findings. In addition, the use of data generated through public health screening activities offers practical insights relevant to TB case-finding in displaced communities.

Several limitations should be considered when interpreting the findings. First, the results may not be generalisable to all displaced populations in Nigeria because the study was conducted in selected locations in Benue State. Secondly, differences in screening coverage, participant selection and access to diagnostic services between displaced and general populations may have influenced the observed proportions.

